# Data Visualization for Epidemiological and Demographic Data for Malaria Surveillance in the Brazilian Amazon, 2007-2019

**DOI:** 10.1101/2021.08.09.21261186

**Authors:** Carlos Eduardo Beluzo, Natália Martins Arruda, Vinícius de Souza Maia, Luciana Correia Alves

## Abstract

Malaria represents one of the main public health problems worldwide and continues to be a great challenge to Brazil which concentrates about 34.4% of the disease cases registered in the American continent. Approximately 99% of Malaria cases occur in Amazonia. In 2017, 194,000 cases were recorded. This increase in the number of cases may be a warning of a possible decline in the effectiveness of control and surveillance programs in the region. The objective of this study is to propose the design of interactive visualizations of data related to Malaria Surveillance in the Brazilian Amazon, between the years of 2007-2019. Data came from SIVEPMalaria. We used data visualization techniques to explore epidemiological and demographic aspects of Malaria Surveillance. We hope tools of this kind can reduce the burden of data extraction and analysis on health staff and local policy makers.

## 1 Introduction

Parasitic diseases affect a large proportion of the world population and exert great influence on the development of many countries. They predominate in less developed regions where the levels of education and basic sanitation are low, causing a high number of deaths (França et al., 2008).

Malaria represents one of the main public health problems worldwide. It is a serious disease caused by parasites that are transmitted to people through the bite of infected female Anopheles mosquitoes and it is present in practically all tropical and subtropical regions of the planet. According to the latest World Malaria report, there were 228 million cases in 2018 compared to 231 million cases in 2017 (WHO, 2019). The estimated number of Malaria deaths stood at 405,000 in 2018, compared with 416,000 deaths in 2017 (WHO, 2020).

Among the species of parasites that cause Malaria in humans, *P. falciparum* and *P. vivax* are the most prevalent. In 2018, *P. falciparum* accounted for 99.7% of estimated cases in the WHO African Region, 50% of cases in the WHO South-East Asia Region, 71% of cases in the Eastern Mediterranean and 65% in the Western Pacific. The WHO African Region continues to carry a disproportionately high share of the disease’s global burden. In 2018, the region was home to 93% of cases and 94% of deaths. The Americas are also at risk. P. vivax is the predominant parasite in the WHO Region of the Americas, representing 75% of cases (WHO, 2019).

The disease continues to be a great challenge to Brazil, that despite of the accumulated knowledge about it and the years of national campaign to fight it, concentrates about 34.4% of the disease cases registered in the American continent (WHO, 2017).

Before 1940, Malaria cases were distributed all over the country. After the execution of successful campaigns and the almost total elimination of the disease in the extra-Amazonian region in the 1950 to 1960, it started concentrating in the Amazonian region (Tauil et al. 1985; Silva e Oliveira, 2002, 2011; Ferreira e Castro, 2016). Approximately 99% of cases in Brazil occur in Amazonia, natural habitat to over 40 species of Anopheles, of which about 20 are potential vectors of the disease (Carlos et al., 2019).

In the last 30 years, there have been fluctuations in the number of cases, with highs of almost 600,000 in 1998–2000 and 2004–2006 (Oliveira-Ferreira et al., 2010). In 2017, 194,000 cases were registered, sounding an alarm to the Malaria control and surveillance program (Jiménez-Muñoz et al., 2016).

Some population groups are at considerably higher risk of contracting Malaria, and developing severe disease, than others. These include infants, children under 5 years of age, pregnant women and patients with HIV/AIDS, as well as non-immune migrants, mobile populations and travellers. Children under 5 years of age are the most vulnerable group affected by it; in 2018, they accounted for 67% (272,000) of all deaths worldwide (WHO, 2019).

Transmission depends on climatic conditions that may affect the number and survival of mosquitoes, such as rainfall patterns, temperature and humidity. In many places, transmission is seasonal, with the peak during and just after the rainy season. Epidemics can occur when climate and other conditions suddenly favour transmission in areas where people have little or no immunity to the disease. They can also occur when people with low immunity move into areas with intense transmission, for instance to find work, or as refugees. Human immunity is another important factor, especially among adults in areas of moderate or intense transmission conditions. Partial immunity is developed over years of exposure, and while it never provides complete protection, it does reduce the risk that an infection will cause severe disease. For this reason, most deaths in Africa occur in young children, whereas in areas with less transmission and low immunity, all age groups are at risk (WHO, 2019).

In Brazil, it should be emphasized that the current organization of the health system and socioeconomic conditions contribute to sustained vulnerability to transmission, since vector control measures are not carried out and adapted according to local needs. Besides, particularly in Amazonia, we can highlight the impact of environmental phenomena that influence the amazon forest climate, as for example, El Niño in the period between 2014-2016, that caused an extension of the extreme drought period, possibly influencing the impressive reduction on the number of cases (Jiménez-Muñoz et al., 2016).

Moreover, migration from States that compose Amazonia, combined with worsening living conditions of migrants, may be partially responsible for an increase in cases observed in recent decades. Importation from neighboring countries may also play an important role in transmission in the Brazilian Amazon. Although the majority of cases in the Brazilian Amazon are autochthonous, case importation between States can provide important insights into disease transmission dynamics. However, human population movement is a key challenge in tackling transmission in Brazil. Whilst temporary movement mostly associated with business, holidays or social visitation, could contribute to small-scale outbreaks in Malaria-free areas, long-term and continuous migration can significantly contribute to the observed and stable transmission dynamics (Carlos et al., 2019).

The majority of diagnosed cases in the extra-Amazonian region are imported mainly from the Amazon region, showing the importance in foreseeing and controlling Malaria in Amazonia and the great influence of people mobility for these areas where medical preparation, training and diagnosis are still insufficient, putting the population in danger of contracting the disease as well as promoting an increase in the number of autochthonous cases (Carlos et al., 2019).

In view of current discussions, a great challenge in fighting Malaria is the sustainability of actions aimed at its control and surveillance. The increase in the number of cases in 2017 in Brazil may be warning of a possible decline in the effectiveness of control and surveillance programs in the region.

At the World Health Assembly in May 2015, WHO launched the Strategy for Malaria elimination in the Greater Mekong subregion (2015–2030), which was endorsed by all the countries in the subregion. Effective surveillance is required at all points on the path to elimination. Stronger surveillance systems are urgently needed to enable a timely and effective response in endemic regions, to prevent outbreaks and resurgences, to track progress, and to hold governments and the global Malaria community accountable (WHO, 2020). Pillar 3 of the Global technical strategy for Malaria 2016–2030 (GTS) is transformation of surveillance into a core intervention in all Malaria-endemic countries and in those countries that have eliminated it but remain susceptible to re-establishment of transmission (WHO, 2019).

Currently, there are few resources for the surveillance of the disease in Amazonia. Usually, they are based on historic data and accomplished by highly capable human sources, making the task costly and frequently unfeasible due to lack of capability to infer knowledge in a timely fashion to accomplish preventive or even corrective action. In addition, the actions that could be taken by public administrators are limited by a lack of tools to explore these data, especially those that are based in data visualization methods, which allow dynamic, interactive analyses.

National Malaria control programs need to take special measures to protect these population groups from infection, taking into consideration their specific circumstances. The Malaria Epidemiological Surveillance System (SIVEP-Malaria) is the main tool used by the National Program for Prevention and Control of Malaria [Programa Nacional de Prevenção e Controle da Malária (PNCM)] for the prevention and control of the disease and to improve quality of information produced about it (Pina-Costa et al., 2014; Wiefels et al., 2016).

Thus, SIVEP-Malaria’s database allows spatial and temporal monitoring of epidemics but also serves to assess the coverage of diagnosis and treatment (Oliveira-Ferreira et al., 2010).

**Aligned with the Sustainable Development Goals and with GTS, and to provide resources to facilitate monitoring, measurement and evaluation of the epidemiological and demographic profile of Malaria, the objective of this study is to propose the design of interactive visualizations of data related to Malaria Surveillance in the Brazilian Amazon, between the years of 2007 and 2019**. In addition, the methods implemented here aim to be important tools to support health managers, since data visualization enables a better understanding of the problem in question, thus favoring the preparation of better services and policies related to surveillance and health care, in this particular case, related to Malaria.

## 2 SIVEP-Malaria

SIVEP-Malaria is a platform for collecting and disseminating important data pertaining to Malaria surveillance in Brazil. The system is an important tool for epidemiological surveillance of Malaria. Much of the information available today about the epidemiological characteristics of the disease originating from SIVEP itself, such as the number of cases, annual parasitic index (IPA), lethality of the disease and the demographic and socioeconomic profile of the patients.

It was implemented in 2003, establishing a distinction between the endemic area - the nine states of the Brazilian Legal Amazon, which concentrates 99% of indigenous cases - and the rest of the national territory (MS, 2019; Carlos et al., 2019). It’s initial implementation had good quality of the epidemiological data related to disease epidemiology, intervention and health status of the patient, however the quality of the data of the other variables took a few years to reach satisfactory levels and some types of variables, such as geographic ones, still have coverage problems (Wiefels et al., 2016). The system is powered by standardized collection instruments. The data are collected in basic units and consolidated by municipalities, then they are consolidated by states and eventually sent to the federal sphere, where they are treated and distributed (MS, 2007).

Malaria cases must be reported both in public and private health systems. In the Amazon region, Malaria is a mandatory regular notification, all cases must be reported within seven days through the system, using the standardized Malaria Case Notification Form. Outside this region, notification is immediate, within 24 hours, by the fastest means available, but the information must be registered in a separate system, the Notifiable Diseases Information System (SINAN - Sistema de Informação de Agravos de Notificação), through the Malaria Investigation Form, therefore, this information is not included in this database.

In the case of Malaria, information is collected about the notifying unit, patient, location of the infection, examination and treatment, when applicable. Here, geographic information of the notifying unit and the patient stands out, which allows analysts to spatialize the incidence of the disease, the demographic and socioeconomic information of the patients, which help to characterize the affected population, and the health information itself, such as the examination and its results, the parasitemia and the treatment regimen used. The data is distributed deidentified, in such a way that it is not possible to identify patients individually.

The data have good coverage and temporal and spatial resolution. The analysis periods available are from 2007 to 2019, with notifications dated by day of occurrence of first symptoms. Geographic information can be disaggregated not only up to the municipality level, but also at the level of the reporting unit (on the health infrastructure side) and locality (on the patient side).

The analysis was carried out for the Brazilian Legal Amazon, which is formed by the following states (UF: Federation Unit): Acre (AC), Amazonas (AM), Amapá (AP), Maranhão (MA), Mato Grosso (MT), Pará (PA), Rondônia (RO), Roraima (RR) and Tocantins (TO). In the case of Maranhão, the area only covers municipalities west of meridian 44.

According to the rules and routines manual MS (2007), the notification flow starts at the notifying unit, by filling in a standardized form numbered in two copies, where the information is recorded the patient and the unit. One of the copies is filed on site, while the other is sent to a typing location in the municipality. The municipality is responsible for gathering the information from the notifying units and sending it to the states, which coordinate the process for all municipalities under their responsibility and then send it to the federal level. A card pre-numbering procedure is used to prevent loss of records.

### 2.1 Dataset

The files are partially available in DBF format, on the DATASUS portal (MS, 2020), through the TABNET system, or through direct requests through the information access law. The dataset built for this study has information from 2007 to 2019. As depicted on Figure 1, our dateset comprises a total of 29,414,406 notified individuals, from Brazillian Legal Amazon, which 12.5% are referent to notified individuals that was than confirmed as infected (positive cases). The rest of them are notified individuals that were not infected.

**Fig. 1.**
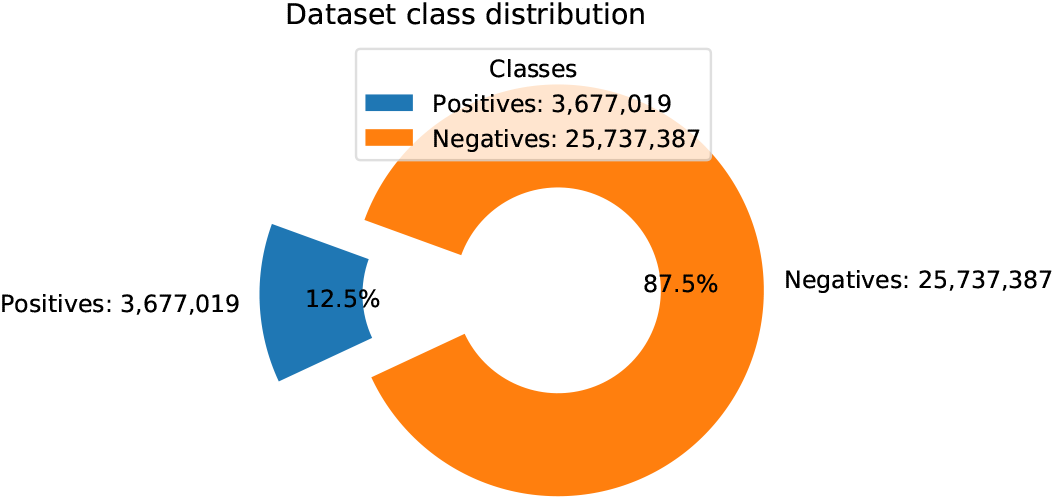
Positive and negative cases distribution among notifications (*Source: SIVEP 2007-2019*).

Each record is uniquely identified by the key fields notification number, notification date and notifying municipality. In total, there are 40 variables that can be classified in three major groups: administrative data, patient information and epidemiology and cure control. The complete list of variables is presented on Table 1.

**Table 1.**
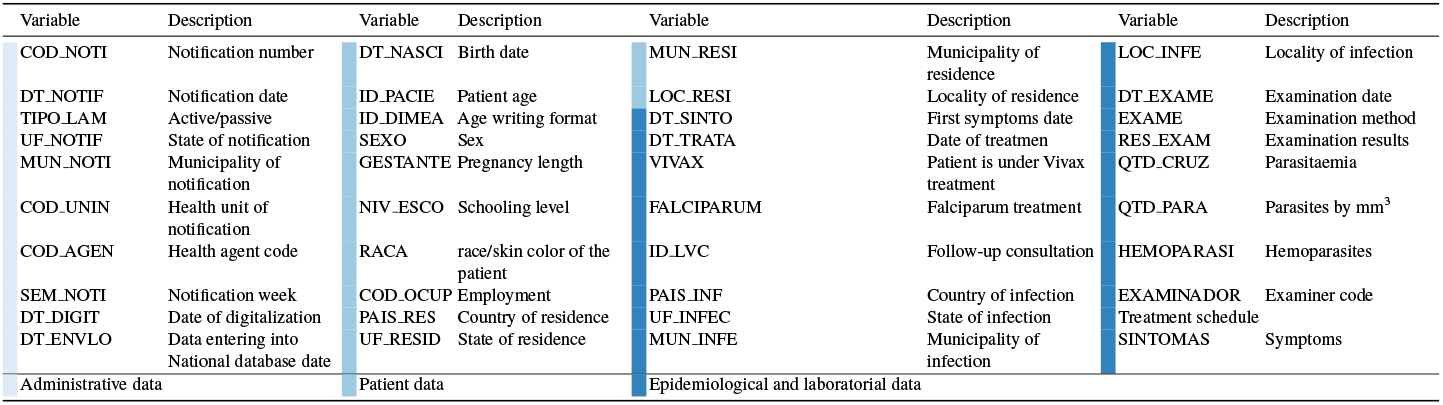
SIVEP data description table - Adapted from WIEFELS et al. (2016)

As reported by Wiefels et al., data availability between variables can be very discrepant, and it is recommended to make a descriptive analysis and exploration to identify for each variable its availability for a specific region and period. Invalid data may occur, but improvements were made in terms of data production and quality over the years. It is possible to mark exactly where and when invalid data occurs and some are concentrated in certain municipalities or localities and also in specific time periods. The outliers and the inconsistencies are not very important statistically, but can be a serious threat to a local study for specific periods, since the accumulation of erroneous data can be precise in space and time (Wiefels et al., 2016).

The dataset used in this study comprises information from 2007 to 2019, and still contains similar concerns regarding data quality. In our sample, considering the entire dataset (i.e., considering the entire period for the whole Legal Amazon region), it was observed that, among the positive individuals, the variables (see Table 1 for descriptions) QTD PARA, NIV ESCO 1, GESTANTE1, ESQUEMA 1, DT NASCI have more than 50% individuals with missing values; and variables VIVAX, RACA, HEMOPARASI, EXAME, EXAMINADOR, FALCIPARUM more than 40%. Some of these contain expected missing values due to form structure and negative case reporting, but a few relevant variables, still have low data quality. Figure 2 depicts the percentage of missing cases for all variables, split by positive and negative cases, after notification.

**Fig. 2.**
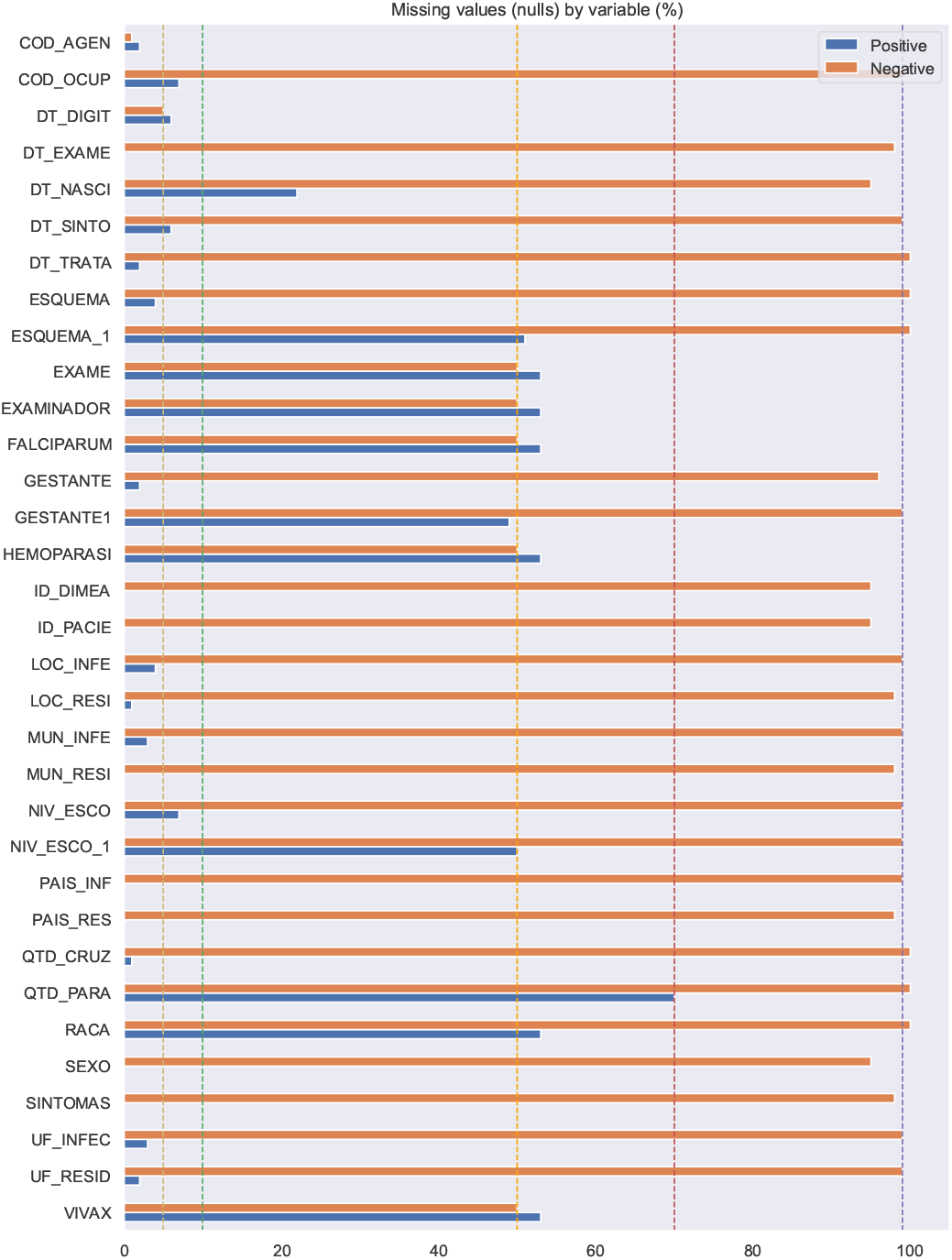
Percentege of null values on each variable on the dataset sample used on this study *Source: SIVEP 2007-2019*.

Also, to calculate the incidence and death rates used in the Fig. 5 we also use population data from Brazilian Institute of Geography and Statistics’ (Instituto Brasileiro de Geografia e Estatística - IBGE) projected population by year, state and age (IBGE, 2020).

### 2.2 Dataset pre-processing

Data visualizations enhance the exploratory and learning phases of both database and collection systems, and guide the methodological adjustment of research. The dataset provides information on the distribution and trends in Malaria and it is a rich data source for both scientific research and health management (Wiefels et al., 2016).

The occurrence of missing or inconsistent data in Brazilian public health repositories is very common, and it mostly happens due to the incorrect filling of handwritten forms. This kind of data inconsistency could lead to missing information and incorrect interpretation of the data visualizations proposed in this paper. In order to tackle this problem the following data pre-processing tasks were performed:

- excluding records reporting age older than 105 years;
- excluding records indicating pregnancies on women younger than 10 years of age;
- excluding records having combined values on variables ID LVC = 2 and TIPO LAM = 3, this is inconsistent because the variable TIPO LAM with value 3 indicates “follow-up appointment”, and variable ID LVC = 2 indicates that “it is not a follow-up appointment”;
- missing value imputation: variables having missing values were treated by year, and filled out with the modal value observed for that variable on that year (see Figure 2 for details).

Considering changes to the forms over the years (inclusion of race/ethnicity, changes in pregnancy, occupation and education categories), as well as information that is not missing, but blank due to the flow of the questionnaire filling procedure, the percentages in figure 2 intend to illustrate overall data quality. For a more detailed description of the issue, see (Wiefels et al., 2016).

## 3 Malaria Data Visualizations

We intend to make tools to orient studies for variable evaluation available. To that end, we have adopted a knowledge discovery approach, focused primarily on obtaining relevant information about each of the two main groups of individuals (positive and negative cases), through data visualization techniques.

As shown on Figure 1, the dataset is unbalanced, when considering, as classes, positive and negative cases (mainly negative). This characteristic of the dataset Figure 3 also appears when data is analyzed at UF level, where the proportion of positive cases among all notifications reaches over 30%, as in PA state. In other regions, such as in MA state, the same proportion is below 10%.

**Fig. 3.**
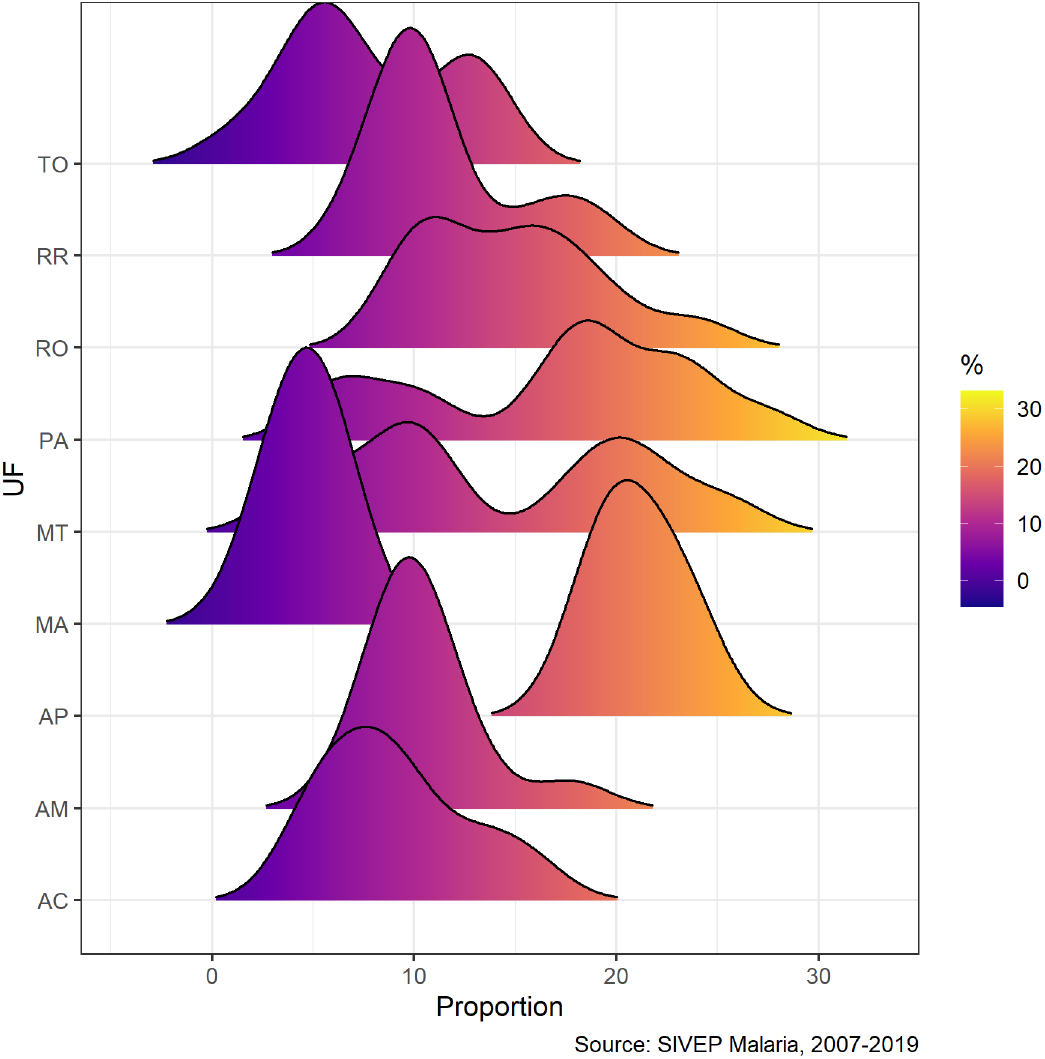
Proportion of positive cases among all investigated cases by state. *Source: SIVEP 2007-2019*.

In this regard, the visualizations presented allow us to explore specific aspects of each of these classes. The visualizations were developed using R (R-Project, 2020) software environment for statistical computing and graphics, specifically implemented with Plotly (Plotly, 2020) graphing library, ggplot2 (Ggplot2, 2020) graphics packages and Shiny package (to build interactive web apps straight from R) (Shiny, 2020). The data was extracted from the pre-processed dataset in comma-separated format and internally summarized as needed through R scripts.

### 3.1 Demographic and socioeconomic profile: Occupation

To build the demographic profile of the population at risk, much information can be extracted and presented on data visualizations, providing public managers with an easier and faster resource for this task. Just to illustrate this potential, the Figure 3 shows changes in distributions of the proportion of positive Malaria cases between 2007 and 2019. Ridgeline plots by state of the Brazilian Amazon indicate a variation in the proportion of cases over the period, with an increase above 20% for the states: AC, MT, PA, RO and RR.

The visualization depicted in Figure 4 consolidates the occupational profile of infected individuals. In this visualization you can choose on the slider a year between 2007 and 2019 and compare on the same page all the states of the Legal Amazon, the distribution of the main activity performed by the patient in the last 15 days according to areas at risk of catching Malaria: Agriculture, Livestock, Domestic, Tourism, Gold Panning, Vegetable Exploration, Hunting/Fishing, Constr. of Roads and Dams, Mining, Traveler, Others and Ignored. This visualization is essential to identify the likely places of infection and to compare the activities between the states of the Legal Amazon to observe the main changes and increases in Malaria infection linked to certain activities.

**Fig. 4.**
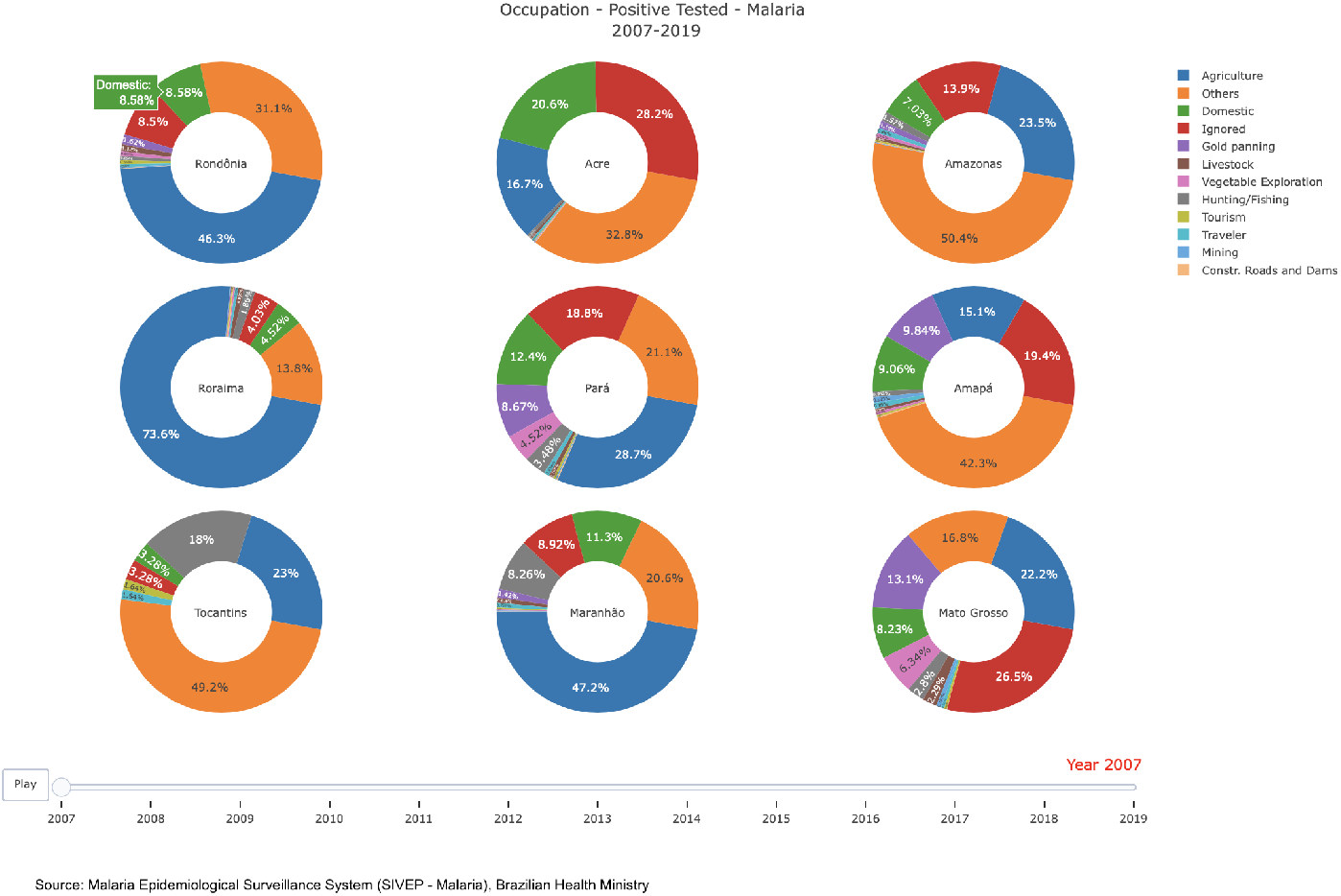
Occupation profile of positive cases data visualization. *Source: SIVEP 2007-2019*. Available at https://arrudanat.shinyapps.io/ocup_eng

For example, in this visualization we can observe the increase in participation of gold mining activity among those tested positive in the state of Pará, between the years 2013 and 2017, reaching 30% of the cases in 2013, whereas in the previous year it was 11% of the cases. The same mining activity in the state of Mato Grosso reached 40% of cases in 2019, compared to 2% in the previous year.

The dataset used on this visualization was a subset of all positive cases of Malaria grouped by year of notification, state of infection and type of occupation.

### 3.2 Malaria incidence rate and death rates

Other important pieces of information are Malaria incidence rates and death rates. This dashboard visualization depicted in Figure 5 shows us such incidence and mortality rates by age group and year of notification and the user can select the state on the side panel. The incidence rate is calculated using the number of new cases divided by the population times 1,000, this index estimates the risk of annual cases and helps analyze geographic and temporal variations in the distribution of the incidence of laboratory positive cases in endemic areas, according to degrees of risk, as part of the set of epidemiological surveillance actions of the disease.

**Fig. 5.**
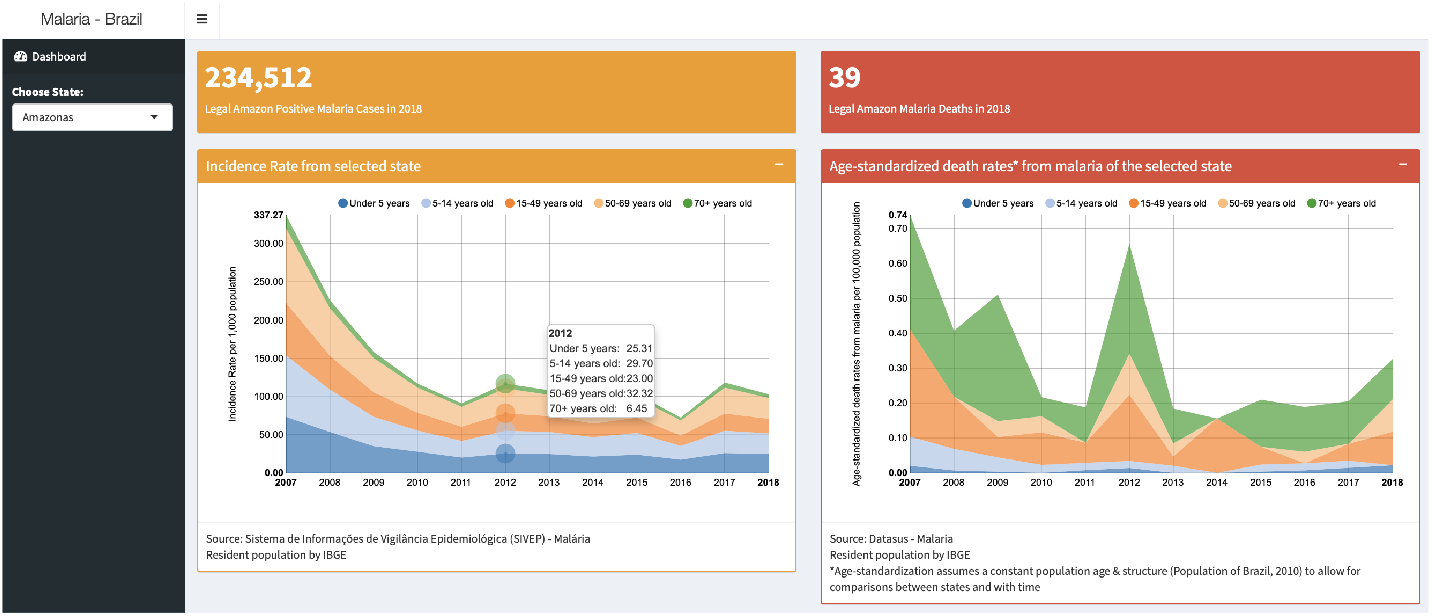
Malaria incidence rate data visualization. *Source: SIVEP 2007-2019*. Available at https://arrudanat.shinyapps.io/taxasMalaria

The other visualization in this dashboard compares Age-standardized death rates from Malaria by 100,000 inhabitants by year, age group and selected state. We use age-standardization death rates to constant population age and structure and allow comparison between states and over time without the effects of changing age distributions across state populations.

This visualization helps us see the age groups at highest risk of infection in a given region and it is possible to compare yearly incidence and states where the death rate is higher side-by-side. We observe in this visualization that Rondônia, Maranhão and Mato Grosso had a decrease in their incidence rate in the period 2007 - 2018 for all age groups, maintaining a level of 2 cases per 1,000 inhabitants. Roraima, in turn, was a state in which there was a big increase in incidence rate between 2016 and 2018. The highest incidence rates are found in the age groups below 5 years and 5 to 14 years in most of the states of the Legal Amazon. However, mortality rates are higher in the age group of 70 or more years old for all states in the period.

The dataset used in this visualization grouped the positive cases of Malaria by age groups (Under 5 years, 5 to 14 years old, 15 to 49 years old, 50 to 69 and 70 years old or more), state and year of notification. To calculate the incidence and death rates we also used data from Brazilian Institute of Geography and Statistics’ (Instituto Brasileiro de Geografia e Estatística - IBGE) projected population by year, state and age (IBGE, 2020).

### 3.3 Distribution between imported and autochthonous cases

Another relevant piece of information is the distribution between imported and autochthonous Malaria cases, as is depicted on Figure 6. On this visualization the viewer can interactively select municipality, period (year) and the variable of interest. The dashboard provides data on the total number of cases in that municipality over the year, a spatial visualization of the sources of important cases (both nationally and internationally) and a selection of charts that compare proportions of autochtochnous and imported cases on several dimensions, such as sex, age, race, occupations, treatment, month of infection, type of parasite, parasite count and treatment regimen prescribed to the patient.

**Fig. 6.**
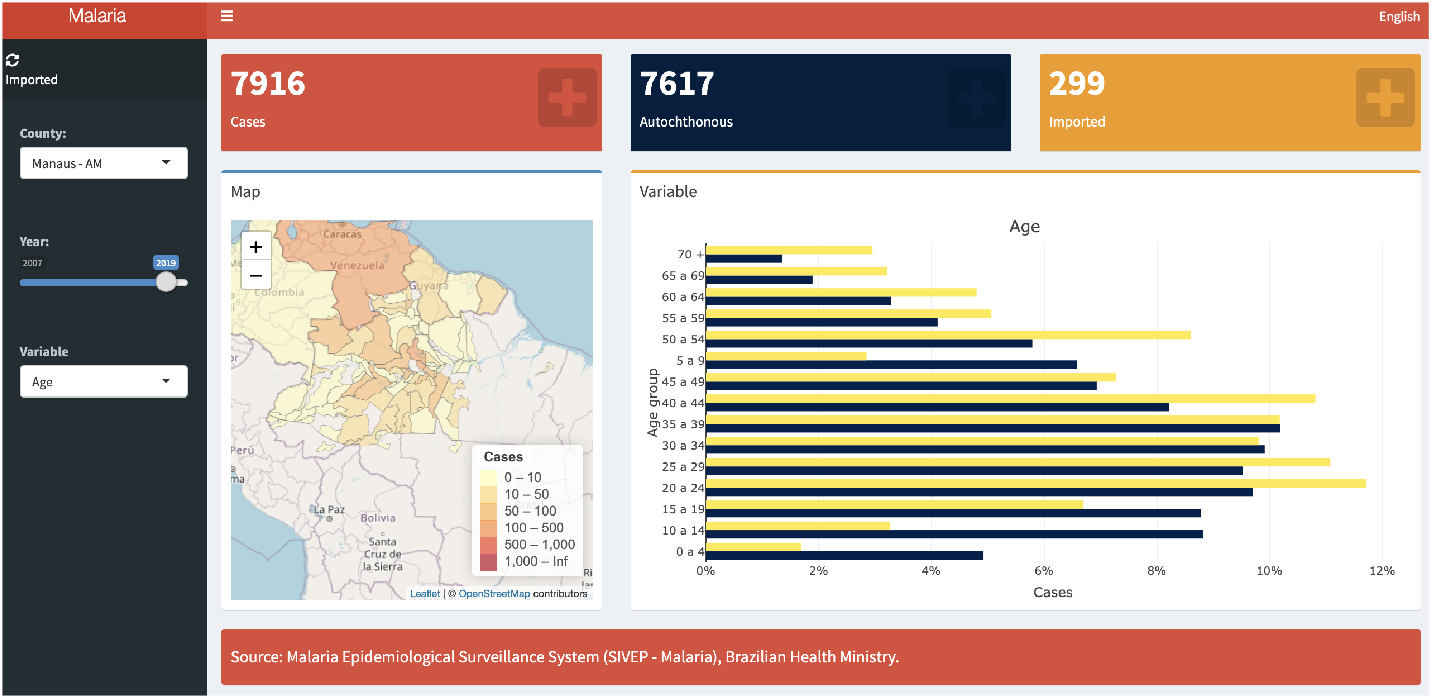
Distribution between imported and autochthonous cases data visualization. *Source: SIVEP 2007-2019*. Available at https://vinicius-maia.shinyapps.io/importadosdash

The goal is to allow non-technical staff to quickly understand the profile of Malaria cases in their local area and allow them to act accordingly in a timely manner. It may also be used as an educational tool, since it allows the viewer to see the evolution of these characteristics over time for a given area. The dataset used for this visualization was the subset of positive cases with valid municipality geospatial data (geocodes from IBGE). The treatment of missing values was the one described in Section 2. Information was summarised to the municipal level, where we used raw case counts and proportions of totals, split by categories of variables of interest where applicable.

Although it was not the main focus behind the application, mapping imported cases allowed us to understand some patterns of case importation. The majority of imported cases in areas where Malaria transmission is still frequent (hundreds or thousands of new cases every year) are locally imported from neighbouring towns and cities. On the other hand, in areas where Malaria transmission is rare, cases are more likely to be imported from further away. When it comes to international importation of cases, there is a small but frequent inflow of cases from neighbouring countries towards frontier towns and also the largest cities, like Manaus and Boa Vista. Despite the existence of these distinct flows, most areas where Malaria incidence is high are fueled primarily by autochthonous and local transmission. The only notable exception to this rule is Boa Vista, capital of the state of Roraima, where flows of imported cases from neighbouring countries like Venezuela and Guyana add to the already significant local case count and even surpass it occasionally, particularly after 2015.

### 3.4 Cases that started treatment within 48 hours and percentage of infections by *Plasmodium falciparum* type of Malaria

It is important to the recovery of patients to begin treatment within 48 hours from the onset of symptoms and to identify regions with high proportion of infections by *Plasmodium Falciparum* (where lethality is higher) and regions with high proportion of infections by *Plasmodium Vivax* (where morbidity is higher). On this visualization depicted in Figure 7 the user can (1) choose a year from the year of notification and (2) select the indicator on the side bar menu.

**Fig. 7.**
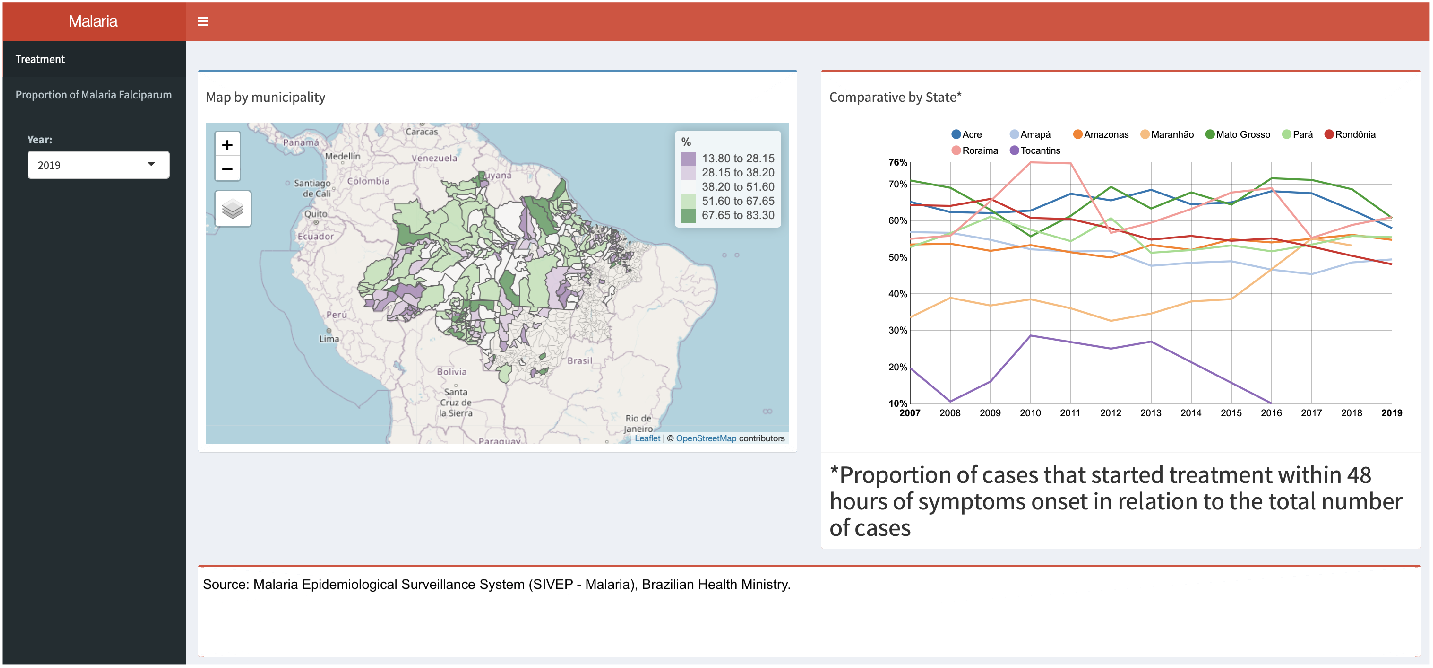
Cases that started treatment within 48 hours. *Source: SIVEP 2007-2019.* Available at https://arrudanat.shinyapps.io/IndicadoresProcesso_eng

We can see three different groups of visualizations on this dashboard. In first group of visualizations we observe a map by municipality that was colored by the proportion of positive cases that started treatment within 48 hours from the first symptoms and a line chart that compares this indicator by time and state. It’s possible from this visualization to see the municipalities that have a low proportion of positive cases, which could indicate inadequate delivery of antimalarial treatment that would contribute to the reduction of disease morbidity and lethality in a specific region.

The other group of visualizations are a map of municipalities colored by percentage of infections by *Plasmodium Falciparum* (responsible for the most serious and lethal forms of the disease) and a line chart comparing periods by state. This visualization helps us see if a particular region has a high percentage of this indicator and tends to indicate poor access to health services and deficient Malaria control and prevention measures. It’s helpful to guide epidemiological surveillance and disease control actions and estimate the quality of prevention and control activities. The last group of visualizations are a map of municipalities color by percentage of infections by *Plasmodium Vivax* (it’s the most common and widespread cause of recurrent Malaria) and a line chart comparing periods by state.

The dataset used on this visualization was a subset of positive cases grouped by year of notification, state, municipalities, *falciparum* category on exam results, *vivax* category and date from the onset symptoms and treatment date all withdrawn from our pre-processed dataset.

Regarding the proportion of treatments that started within two days of the onset of symptoms, the state of Amazonas maintained the proportion of approximately 50% during the entire period of 2007-2019. The State of Acre, in turn, maintains an average of 60%. In relation to the indicator that shows the proportion of the most serious type of the disease, Malaria *Falciparum*, the map indicates that few municipalities have a high proportion of Malaria *Falciparum* and most municipalities are in the state of Mato Grosso in the period from 2007 to 2019. The state of Acre has the highest proportion between the years 2013 to 2019 in relation to all other states in the Legal Amazon. Comparatively, looking at the proportion of cases by Malaria Vivax, Acre has the lowest levels, around 70% of cases being of the vivax type, against almost 93% in Amapá.

## 4 Conclusion

Malaria is a major concern of public health in the world, and continues to be a great challenge to Brazil. After years of successful campaigns, it was almost fully eliminated in the extra-Amazonian region, and is concentrated in the Amazonian region.

Some population groups are at higher risk, including infants, pregnant women and patients with HIV/AIDS, mobile populations and travellers, as such, data analyses tools are required to better understand the problem. Currently, there are few such resources for the surveillance of cases in Amazonia. Following the Sustainable Development Goals and with GTS defined by WHO, this study provided data visualizations as a tool to contribute to Malaria Surveillance in the Brazilian Amazon.

The findings of this study are encouraging. We presented as results of this paper four interactive and one static data visualizations that allow data exploration of a few aspects of the SIVEP-Malaria dataset: (1) a visualization that consolidates the occupational profile of the infected individuals to illustrate the potential for the development of demographic and socioeconomic data visualizations; (2) Data visualization to explore malaria incidence rate and death rates, by Federative Unit (State), by age group and year of notification; (3) Data visualization to explore distribution between imported and autochthonous cases, by municipality and year; and (4) Data visualization to explore cases that started treatment within 48 hours and percentage of infections by *Plasmodium falciparum* type of malaria, by year of notification.

The World Health Organization continues to highlight the urgent need for new and improved tools in the global response to Malaria. Surveillance is therefore the basis of operational activities in settings of any level of transmission. Its objective is to support reduction of the burden of the disease, eliminate it and prevent its re-establishment. We hope tools of this kind can reduce the barrier to data use in policy decisions as it reduces the burden of data extraction and analysis on health staff and local policy makers.

## Data Availability

SIVEP-Malaria dataset was made available upon request for access to information to the Brazilian Ministry of Health, in accordance with Law No. 12,527, of November 18, 2011 (L12527).

## Statement

This work uses data in the public domain, made available upon request for access to information to the Brazilian Ministry of Health, in accordance with Law No. 12,527, of November 18, 2011 (L12527). The data is de-identified, and the results present aggregated information, therefore, this work is exempt from being evaluated by the Ethics and Research Committee, according to Resolution No. 510, of April 7, 2016, of the National Health Council (Resolution CNS/MS 510/16).

## Acknowledgements

Acknowledgment

We thank the Bill & Melinda Gates Foundation (Process ID INV 003970) and Brazilian Ministry of Health through the National Council for Scientific and Technological Development (CNPq) (Process 443048/2019-3) by financial support.

## External Resources

The visualizations can be accessed using QR Codes bellow.

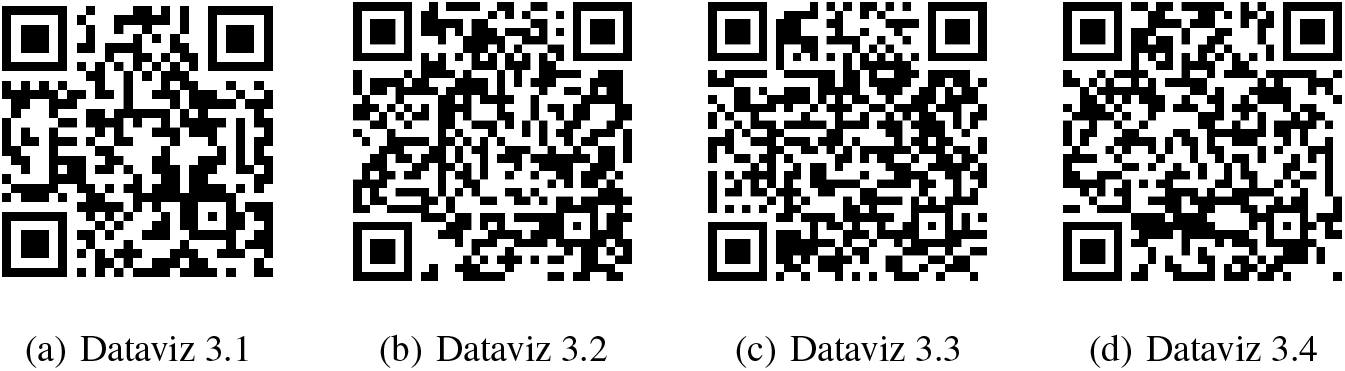

